# The ClinGen Syndromic Disorders Gene Curation Expert Panel: Assessing the Clinical Validity of 111 Gene-Disease Relationships

**DOI:** 10.1101/2024.11.19.24317561

**Authors:** Eleanor Broeren, Vanessa Gitau, Alicia Byrne, Pamela Ajuyah, Marie Balzotti, Jonathan Berg, Krista Bluske, B. Monica Bowen, Matthew P. Brown, Amanda Buchanan, Brendan Burns, Nicole J Burns, Anjana Chandrasekhar, Aditi Chawla, Jessica Chong, Maya Chopra, Amanda Clause, Marina DiStefano, Stephanie DiTroia, Marwa Elnagheeb, Amanda Girod, Himanshu Goel, Katie Golden-Grant, Thuong Ha, Ada Hamosh, Jennifer Huang, Madeline Hughes, Saumya Jamuar, Sylvia Kam, Akanchha Kesari, Ai Ling Koh, Rhonda Lassiter, Sarah Leigh, Gabrielle Lemire, Jiin Ying Lim, Alka Malhotra, Hannah McCurry, Becky Milewski, Shahida Moosa, Stephen Murray, Emma Owens, Emma Palmer, Brooke Palus, Mayher Patel, Revathi Rajkumar, Julie Ratliff, F. Lucy Raymond, Bruno Della Ripa Rodrigues Assis, Samin Sajan, Zinayida Schlachetzki, Sarah Schmidt, Zornitza Stark, Samuel Strom, Julie Taylor, Courtney Thaxton, Devon Thrush, Sabrina Toro, Kezang Tshering, Nicole Vasilevsky, Bess Wayburn, Ryan Webb, Anne O’Donnell-Luria, Alison J. Coffey

## Abstract

**Purpose:** The Clinical Genome Resource (ClinGen) Gene Curation Expert Panels (GCEPs) have historically focused on specific organ systems or phenotypes; thus, the ClinGen Syndromic Disorders GCEP (SD-GCEP) was formed to address an unmet need.

**Methods:** The SD-GCEP applied ClinGen’s framework to evaluate the clinical validity of genes associated with rare syndromic disorders. 111 Gene-Disease Relationships (GDRs) associated with 100 genes spanning the clinical spectrum of syndromic disorders were curated.

**Results:** From April 2020 through March 2024, 38 precurations were performed on genes with multiple disease relationships and were reviewed to determine if the disorders were part of a spectrum or distinct entities. 14 genes were lumped into a single disease entity and 24 were split into separate entities, of which 11 were curated by the SD-GCEP. A full review of 111 GDRs for 100 genes followed, with 78 classified as Definitive, 9 as Strong, 15 as Moderate, and 9 as Limited highlighting where further data are needed. All diseases involved two or more organ systems, while the majority (88/111 GDRs, 79.2%) had five or more organ systems affected.

**Conclusion:** The SD-GCEP addresses a critical gap in gene curation efforts, enabling inclusion of genes for syndromic disorders in clinical testing and contributing to keeping pace with the rapid discovery of new genetic syndromes.

## Introduction

Syndromic disorders are complex conditions involving multiple organs or body systems and are highly enriched among infants with structural birth defects. Combined, genetic disorders and congenital malformations represent the leading cause of infant mortality in the US, responsible for 20% of deaths,^1,2^ and have a large health economic burden in children and adults.^3^ Syndromic disorders are one of the largest and fastest growing categories of disorders with multiple new syndromes published every month.^4^ To ensure the clinical validity of genetic testing for syndromic disorders, it is critical to rigorously evaluate the underlying evidence and classify the validity of purported gene-disease relationships (GDRs).

GDR validity curation is an essential first step for accurate and consistent clinical interpretation across variant triage, classification, and reporting.^5^ The American College of Medical Genetics and Genomics (ACMG) recommends that diagnostic gene panels include only GDRs that meet the Clinical Genome Resource’s (ClinGen) criteria for Definitive, Strong, or Moderate evidence. GDRs with a classification of Limited should typically only be assessed as part of exploratory exome or genome analysis, while GDRs classified as Disputed or Refuted are not appropriate for diagnostic testing.^6^ For variant classification,^7^ variants found in genes with only a Moderate GDR should not exceed a classification of likely pathogenic, and variants in genes with Limited evidence should not be classified above a variant of uncertain significance.^6^

The Clinical Genome Resource (ClinGen), a National Institutes of Health (NIH) National Human Genome Research Institute (NHGRI)-funded initiative, is building an authoritative central resource to define the clinical relevance of genes and variants for use in precision medicine and research.^8^ To achieve this goal, ClinGen has developed an evidence-based gene-disease validity curation framework that allows semiquantitative assessment of the strength of evidence underlying GDRs which is then translated into seven qualitative classifications. Four classification categories indicate evidence supporting a GDR (Definitive, Strong, Moderate, and Limited) while two categories indicate contradictory evidence (Disputed, Refuted). A Strong classification has the same level of evidence as a Definitive classification, but less than three years have passed between publications documenting human genetic evidence. No Known Disease Relationship indicates the GDR is not supported by human genetic evidence. Curated evidence is reviewed by gene curation expert panels (GCEPs), with appropriate clinical and laboratory expertise, and the resultant classification confirmed or adjusted based on expert insight.^9^

ClinGen GCEPs have historically focused on a specific organ system or phenotype. The challenge of the growing number of syndromic disorders published each year not covered by these curation efforts provided an opportunity for a GCEP to take a different approach to address this unmet need. The ClinGen Syndromic Disorders Gene Curation Expert Panel (SD-GCEP; https://clinicalgenome.org/affiliation/40060) was therefore established in March 2020 to classify the clinical validity of GDRs involving multiple body systems not under evaluation by another GCEP and is co-funded by the National Institute of Child Health and Human Development (NICHD) and National Institute of Neurological Disorders and Stroke (NINDS). The SD-GCEP is currently composed of 46 expert and biocurator members (and 25 former members) across 41 institutions and 6 continents. The GCEP meets twice a month plus additional quarterly meetings to accommodate as many time zones as possible. Here, we describe the work of the SD-GCEP, providing the community with outcomes and updates from the group.

## Methods

### Membership

The membership of the SD-GCEP is composed of medical geneticists, genetic counselors, clinical molecular geneticists, variant scientists, and basic scientists, as well as staff biocurators from ClinGen. These members are largely volunteers from academic institutes, clinical laboratories, and organizations that provide online gene-level resources. Initial membership was solicited through invitation or self-nomination. New members can volunteer through ClinGen’s Community Curation Database (https://ccdb.clinicalgenome.org/apply) or by reaching out to the SD-GCEP coordinator directly (https://clinicalgenome.org/affiliation/40060).

### Identifying Relevant Genes

Five approaches have been used during the existence of the SD-GCEP to identify relevant GDRs for curation. Approach 1 was to identify syndromic GDRs most frequently tested in clinical laboratories, prioritized by the number of tests in the Genetic Testing Registry (GTR)^10^ and the number of pathogenic or likely pathogenic variants in the gene in ClinVar. Approach 2 identified GDRs detected through clinical genome or exome sequencing performed by diagnostic laboratories within the membership of the SD-GCEP, utilizing and building upon the curation efforts performed as part of this testing. Approach 3 identified new GDRs from research consortia including NHGRI’s Centers of Mendelian Genomics and GREGoR consortium, again utilizing existing internal efforts. Approach 4 included any GDRs requested by other GCEPs for phenotypes requiring the broad expertise of the SD-GCEP. Most recently, Approach 5 was used to capture additional syndromic GDRs that have been curated by groups not using the ClinGen framework by first searching the Gene Curation Coalition (GenCC) database (https://search.thegencc.org/) for disease assertions with “syndrome” in name, then prioritizing Strong or Definitive classifications given these are often included on gene panels. The genes from all approaches were reviewed to ensure the disease assertions were syndromic in nature, rather than pertaining to a single organ system, and the final list reviewed and approved by the SD-GCEP chairs.

### Precuration for Genes with Multiple Disease Assertions

ClinGen’s precuration is the process of evaluating available information for GDRs to determine the disease entity and mode of inheritance to be curated.^11^ When evaluating genes with multiple disease assertions listed within ontologies such as Online Mendelian Inheritance in Man (OMIM),^12^ Orphanet,^13^ Mondo Disease Ontology (Mondo),^14^ or the literature, the SD-GCEP refers to ClinGen’s Lumping and Splitting guidelines^15^ to precurate the gene. ‘Lumping’ involves combining two or more conditions into a single disease entity, while ‘splitting’ involves separating disease assertions into distinct entities. To inform this decision, the molecular mechanism, phenotypic variability within and across families, and the mode of inheritance are reviewed for each disease assertion, and the decision to lump or split is then voted on by the expert panel. Following ClinGen’s guidelines, disorders are generally lumped into a single entity when the underlying molecular mechanism is consistent, and the clinical features represent a spectrum of the same condition. When there is insufficient data on the molecular mechanism or uncertainty on the phenotypic spectrum, disadvantages of splitting include dilution of evidence across multiple GDRs and underweighting of the final curation strength. The expert panel decides on the most appropriate name for the entity to be curated by applying the dyadic naming convention(s) for lumped conditions, which are defined by the ClinGen Guidance and Recommendations for Monogenic Disease Nomenclature.^16^ The naming decisions are communicated to OMIM, Mondo, and Orphanet for consideration of inclusion in ontology.

### Curation and Expert Panel Review

The SD-GCEP evaluates the clinical validity of GDRs according to ClinGen’s gene curation workflow,^9^ and curation is performed by SD-GCEP curators using ClinGen’s Gene Curation Interface (GCI)^17^. Genetic and experimental data, either supporting or disputing the validity of the disease relationship, are examined and classified using the current standard operating procedures document (for this study, versions 7 to 10 depending on the date of curation (https://clinicalgenome.org/docs/?doc-type=curation-activity-procedures&curation-procedure=gene-disease-validity)) by a SD-GCEP biocurator. On average, two biocurators each present one curation per meeting on either the biweekly or quarterly video conference call. The final decision is voted on and approved by the members in attendance at the meeting, including both biocurators and experts. Curators submit evidence summaries for approval by the chairs which are then published to the ClinGen website for broad community access.

### SD-GCEP Specific Guidelines

Many GCEPs adopt internal modifications to the ClinGen Gene-Disease Validity Standard Operating Procedures in places where there is a degree of subjectivity. In addition to the formal guidelines, and to help with consistency, the SD-GCEP has created a comprehensive “SD-GCEP Scoring Guidance and Specifications Document” and a short form that includes additional guidance and detailed information specific to the SD-GCEP, including where to look for data and how to score them (Supplemental Material). Of note, considering human genetic evidence, the SD-GCEP specifically downgrades loss-of-function variants for homozygosity and if consanguinity cannot be ruled out, because these individuals are likely have multiple homozygous variants due to runs of homozygosity, and it may be unclear which homozygous variants are causative^18^ (Table 1). Due to the rarity of many syndromic phenotypes meaning the number of unique pathogenic variants can be limited, the SD-GCEP allows scoring of multiple observations of the same variant when *de novo* occurrence is proven. Each observation is awarded the default score for the variant type as well as the upgrade for *de novo* status. Recurrently occurring variants that present with a highly specific phenotype and are known to act via a dominant mechanism are upgraded (Table 1). In considering experimental evidence, non-human animal models are thoroughly evaluated to determine whether the phenotype appropriately recapitulates the syndromic human disease. Models that lack reproducibility of multiple human phenotypes are downgraded to 1 point, instead of the default 2 points (Table 1). Models where a human disease variant has been knocked-in are considered for the maximum 4 points. Morpholino-mediated knock-down models are downgraded to 1 point, unless re-expression of the wild-type allele demonstrates rescue of the phenotype.^19^

**Table 1.**
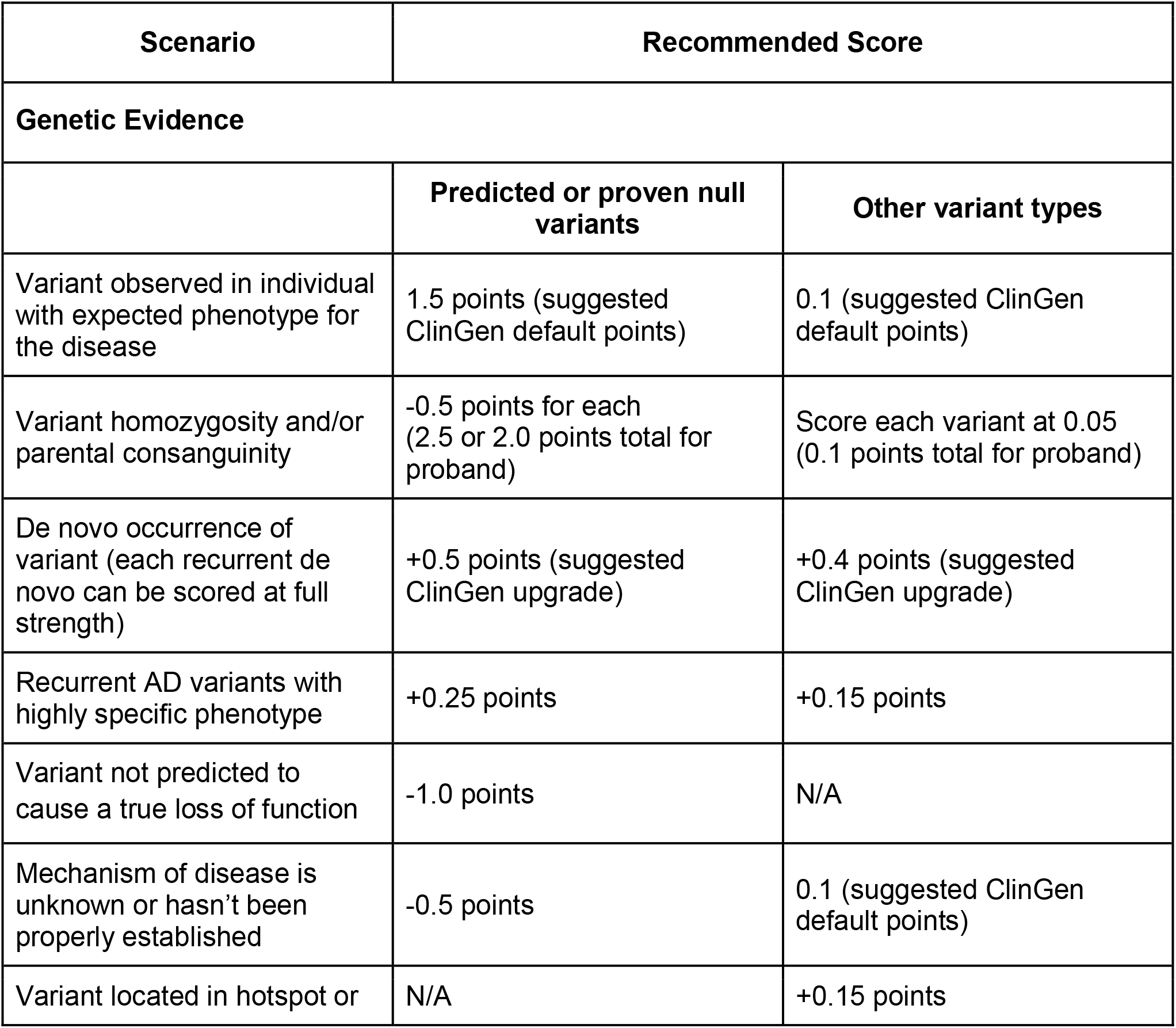
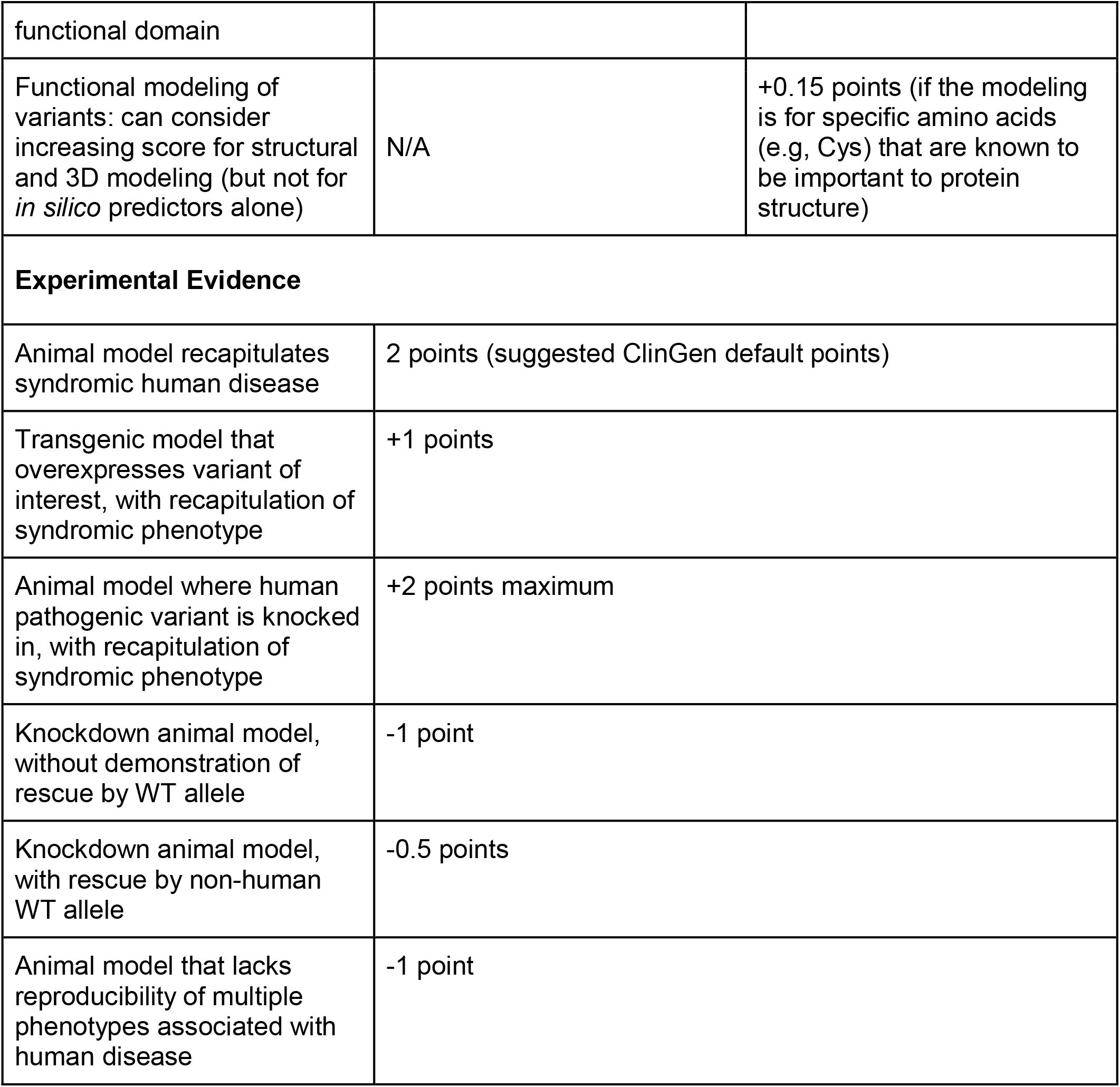
SD-GCEP scoring guidance.

## Results

### Assessing Gene-Disease Validity Across Syndromic Disorders

In the first four years, from April 2020 through March 2024, the SD-GCEP performed 38 precurations and 111 curations of GDRs involving 100 genes. For precurations, 14 genes with multiple assertions were lumped and curated as a single entity and 24 were split. For curations, 78 GDRs were classified as Definitive, 9 as Strong, 15 as Moderate, and 9 as Limited (Figure 1A+B). For the 111 GDRs, 60 had an autosomal recessive (AR) mode of inheritance, 45 were autosomal dominant (AD), three were X-linked recessive, two were X-linked dominant, and one had an unclear mode of inheritance (Figure 1C). The unclear mode of inheritance is associated with the GDR for *UNC13A* and congenital nervous system disorder, which was curated by the SD-GCEP in 2021. At the time, three variants including missense and stop-gained variants had been reported in association with a neurodevelopmental syndrome characterized by variable features of developmental delay, seizures, microcephaly, and myopathy/movement disorders.^20– 22^ Based on the limited number of reported cases, a distinct mode of inheritance, mechanism of disease, and phenotypic spectrum could not be determined,therefore the classification remains limited and provisional until further evidence is published.

**Figure 1.**
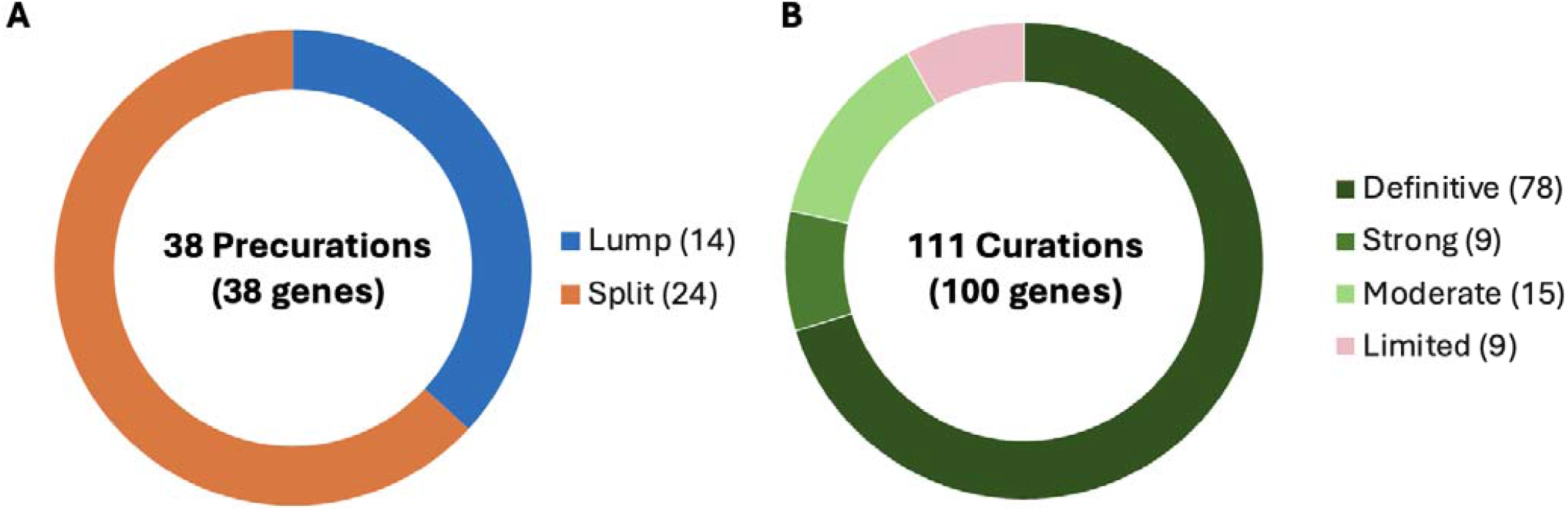
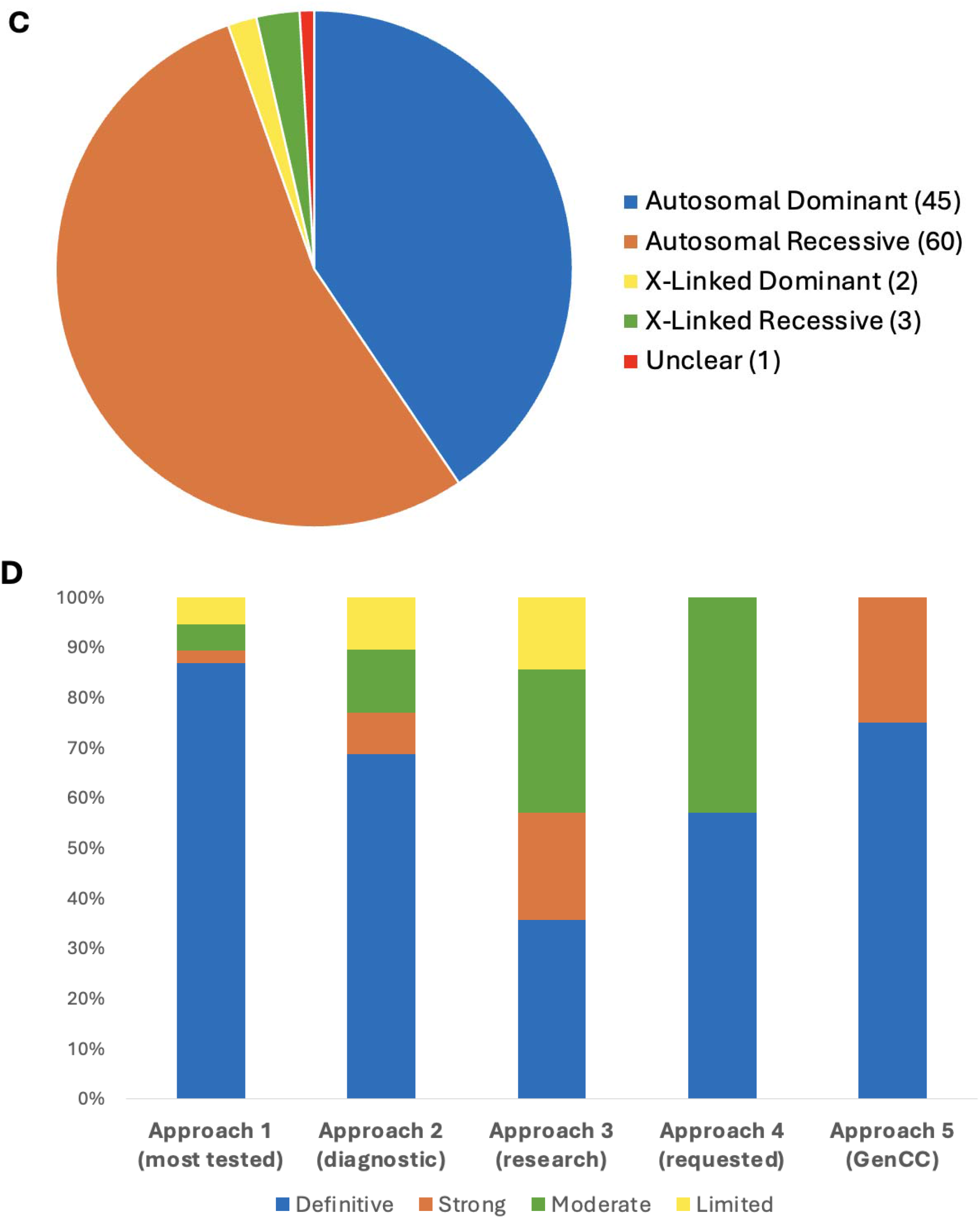
Precurations and curations performed from April 2020 - March 2024. **A)** Summary of lumping and splitting decisions for the 38 precurations performed to date. **(B)** Summary of the final classifications for the 111 approved GDR classifications to date. **(C)** Summary of mode of inheritance of the 111 approved GDRs to date. **(D)** Summary of final classifications for GDRs across the five curation approaches. Approach 1 (38 genes - GDRs most frequently tested in clinical laboratories): 33 Definitive, 1 Strong, 2 Moderate, 2 Limited; Approach 2 (47 genes - GDRs through clinical genome or exome sequencing performed by diagnostic laboratories within the membership of the SD-GCEP): 33 Definitive, 4 Strong, 6 Moderate, 5 Limited; Approach 3 (14 genes - new GDRs from research consortia including NHGRI’s Centers of Mendelian Genomics and GREGoR consortium): 5 Definitive, 3 Strong, 4 Moderate, 2 Limited; Approach 4 (7 genes - GDRs requested by other GCEPs for phenotypes requiring the broad expertise of the SD-GCEP): 4 Definitive, 3 Moderate; Approach 5 (4 genes as of March 2024 - syndromic GDRs in GenCC with Strong or Definitive classifications not curated by other GCEPs): 3 Definitive, 1 Strong.

The 38 GDR curations from the most frequently tested genes in clinical laboratories were more often classified as Definitive (“Approach 1”: 33 Definitive, 1 Strong, 2 Moderate, 2 Limited). This was similar to the distribution seen for the 48 GDR curations identified through clinical genomic sequencing performed by diagnostic laboratories by the membership of the SD-GCEP (“Approach 2”: 33 Definitive, 4 Strong, 6 Moderate, 5 Limited). A broader distribution was seen for the 14 GDR curations from within a research setting (“Approach 3”: 5 Definitive, 3 Strong, 4 Moderate, 2 Limited). Seven GDR curations came from collaboration with other GCEPs (Approach 4”: 4 Definitive, 3 Moderate), and 4 GDR curations came from the more recently designed gene list looking at Strong and Definitive GDRs in the GenCC that have not yet been curated by ClinGen (“Approach “5: 3 Definitive, 1 Strong) (Figure 1D). It was an open question whether GDRs without animal models would have sufficient evidence to reach classifications necessary to be included in diagnostic testing panels (Moderate or above); however, of the curations for which there were no animal models available to score, 8 were classified as Definitive, 4 as Strong, 4 as Moderate, and only 4 as Limited (Figure 2A). For example, the GDR for *ARSL* and X-linked chondrodysplasia punctata 1 is well defined in the literature, with variants in at least 50 probands in seven publications scored in this curation ^23–29^, maxing out the genetic evidence at 12 points and reaching a Definitive classification, but has no model organism. Overall, the presence of an animal model did correlate with the curation classification. 86.1% of curations classified as Definitive, 55.6% of curations classified as Strong, 66.7% of curations classified as Moderate, and 55.6% of curations classified as Limited had an animal model scored. Mouse models were the predominant model organism scored, while zebrafish, *Xenopus, Drosophila, C. elegans*, and other models have also been scored (Figure 2B). 82% of all curations had animal models counted as experimental data (Figure 2C).

**Figure 2.**
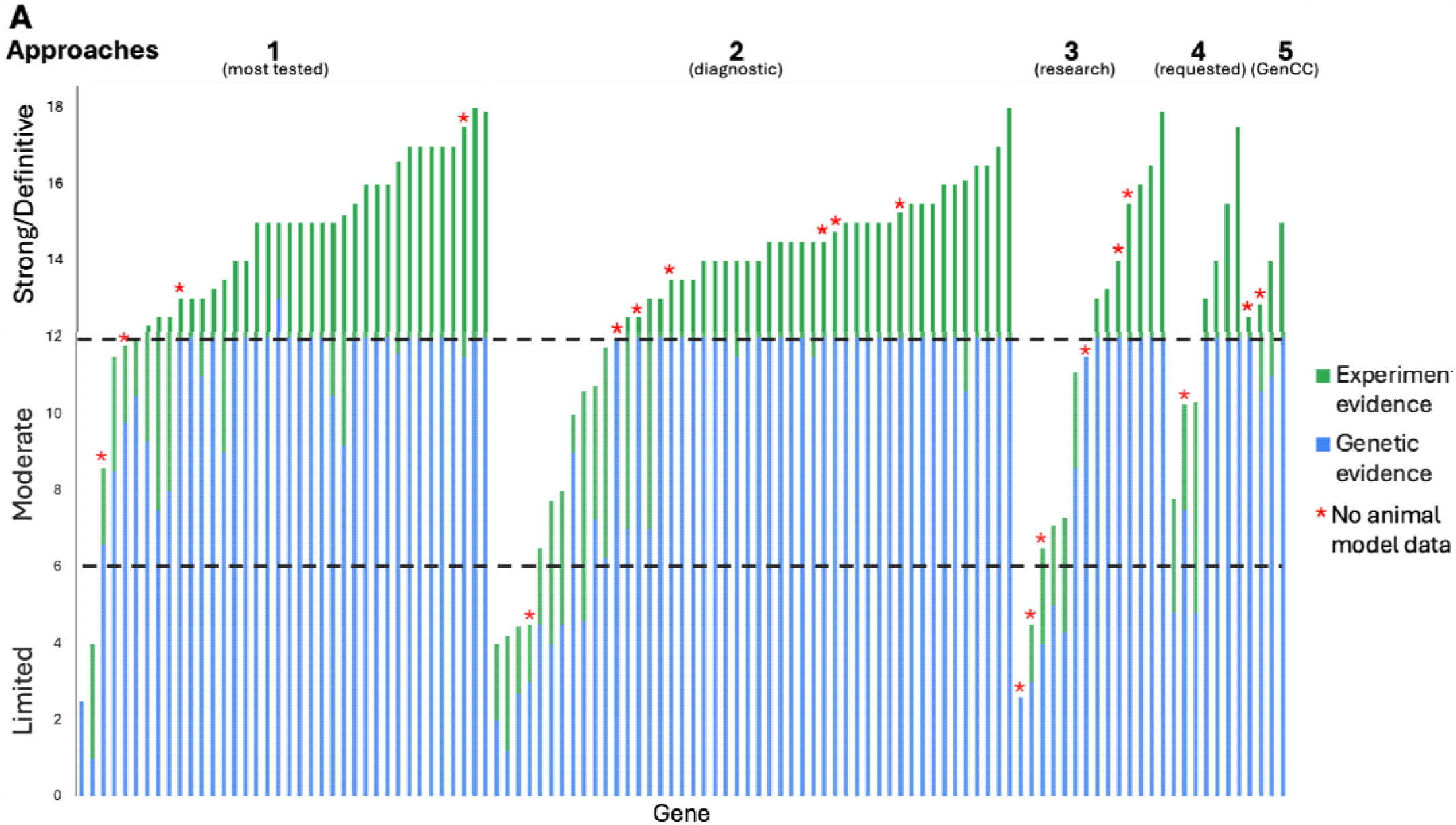
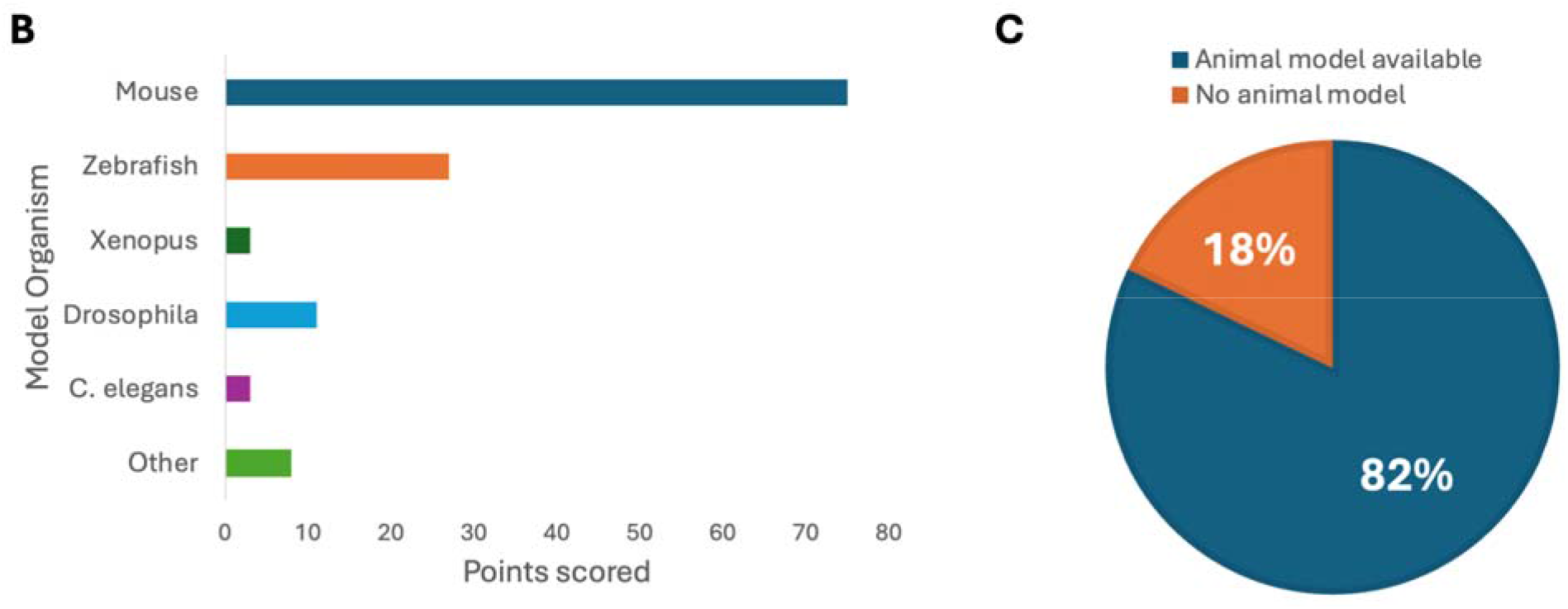
SD-GCEP Curation Evidence. (A) The number of points awarded for genetic and experimental evidence for each gene represented by the bar height (blue = genetic evidence; green = experimental evidence). GDRs where no animal model was available are starred. (B) The majority of genes (82%) had an animal model with 92% of sufficient quality and overlap with the human phenotype to be scored in the ClinGen curation framework. (C) Mouse models were the predominant model organism scored.

### Phenotypic Diversity in Syndromic Disorders

GDRs curated by the SD-GCEP are syndromic in nature, demonstrated by the diseases involved affecting more than one organ system. The number of HPO terms under each of the 23 top-level terms by organ system (direct descendants of “Phenotypic abnormality, “HPO:0000118”) for each of the disease assertions for GDRs under curation was counted (Figure 3). All of the 23 different organ systems were affected in at least one of the curated GDRs. Of the 111 GDRs curated, a median of 8 organ systems were affected per disease. All diseases involved two or more organ systems, while the majority (88/111 GDRs, 79.2%) had five or more organ systems affected (Figure 3). The most commonly involved organ systems included the nervous system, head and neck, eye, and the skeletal system.

**Figure 3.**
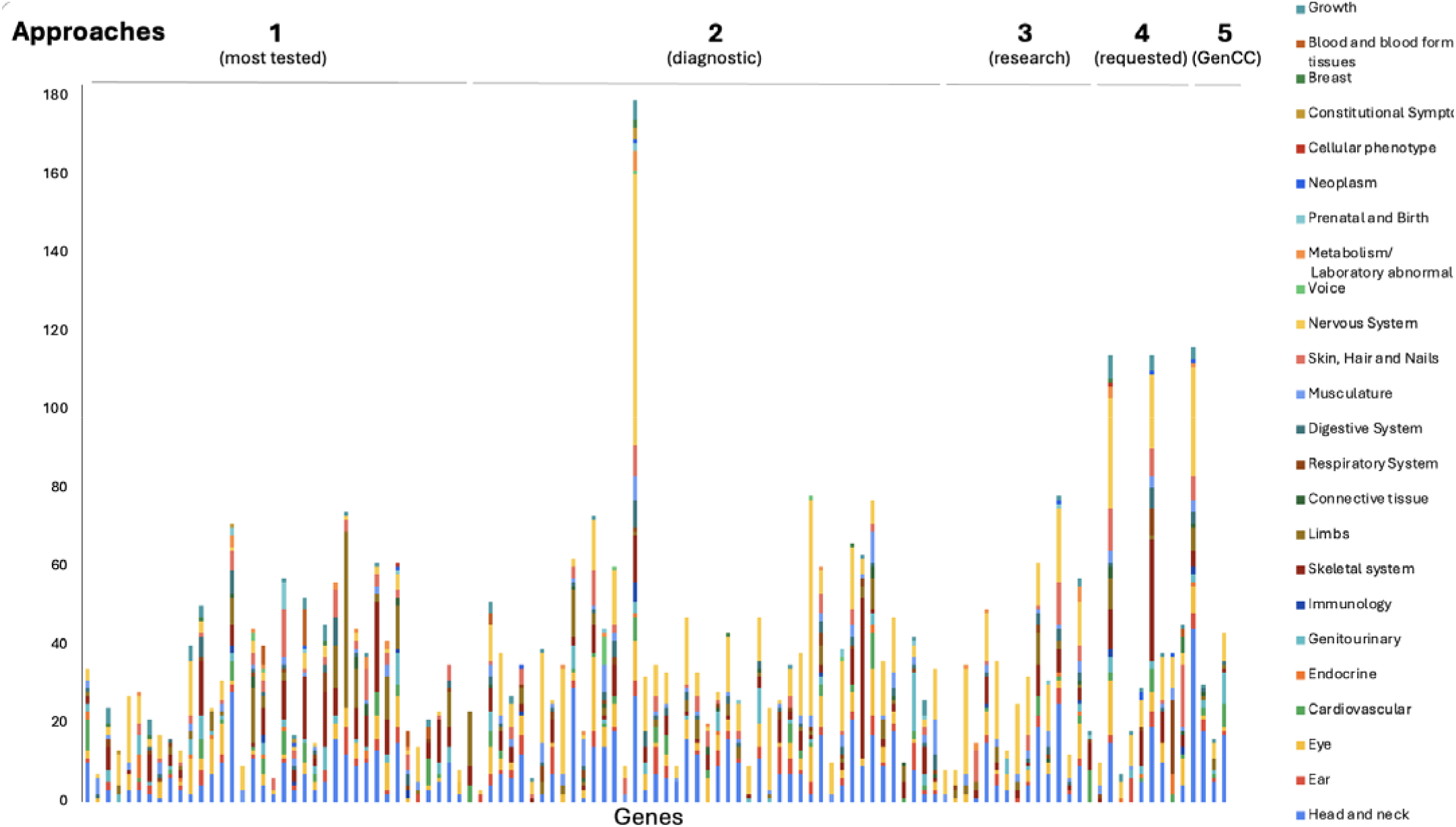
GDRs curated are highly syndromic in nature. HPO higher order terms representing the phenotypic features associated with each disease assertion were graphed by counting the number of HPO terms under each higher order term. For terms that were lumped, HPO terms for all of the assertions were combined and counted. All diseases involved two or more organ systems, while the majority (88/111 GDRs, 79.2%) had five or more organ systems affected.

### Collaboration Between GCEPs

The overlapping phenotypic features between syndromic disorders and other disease areas necessitated communication and collaboration between the SD-GCEP and other GCEPs.

Genes are transferred to different GCEPs based on the specific expertise needed. For example, if a disease assertion is more syndromic than a GCEP is comfortable reviewing, they may transfer the gene to be curated by the SD-GCEP. Conversely, if a disease assertion originally thought to be syndromic mostly affects one organ system, it will be transferred to another GCEP if there is a specific GCEP for that disease area. Transfer to a more suitable GCEP also happened when new GCEPs were started after the SD-GCEP planned GDR lists were composed. Over the past four years, the SD-GCEP has collaborated and exchanged genes with eight GCEPs: <u>Cerebral Palsy</u>, <u>Congenital Myopathies</u>, <u>Craniofacial Malformations</u>, <u>Glomerulopathy</u>, <u>Intellectual Disability and Autism</u>, <u>Kidney Cystic and Ciliopathy Disorders</u>, <u>Retina</u>, and <u>Severe Combined Immune Deficiency and Combined Immune Deficiency</u> GCEPs (Figure 4). Through this collaboration, 10 genes have been shared between the SD-GCEP and another GCEP (*USP7, NFIX, GATAD2B, ASHL1, SMO, LMX1B, IKBKG, INPP5E, CPLANE1, TCTN2*). For example, the GDR *CPLANE1* and Joubert syndrome 17 was shared between the SD-GCEP from the Kidney Cystic and Ciliopathy Disorders (KCCD) GCEP due to overlapping phenotypes. The KCCD GCEP has curated several forms of Joubert syndrome because of the renal manifestations; however, patients with Joubert syndrome 17 and variants in *CPLANE1* were not found to have kidney involvement,^30^ so the SD-GCEP and KCCD GCEPs shared the curation and both are acknowledged as contributors on the ClinGen website. Additionally, eight genes have been transferred from the SD-GCEP to another GCEP (*KMT2E, MED13, TANC2, MKKS, MUSK, DOK7, GFPT, CHRND*), and nine genes have been transferred from another GCEP to the SD-GCEP (*HCCS, MYO5A, BCOR, EP300, SOX3, PNPLA6, NKX2-1, C19ORF12, ZNF423*).

**Figure 4.**
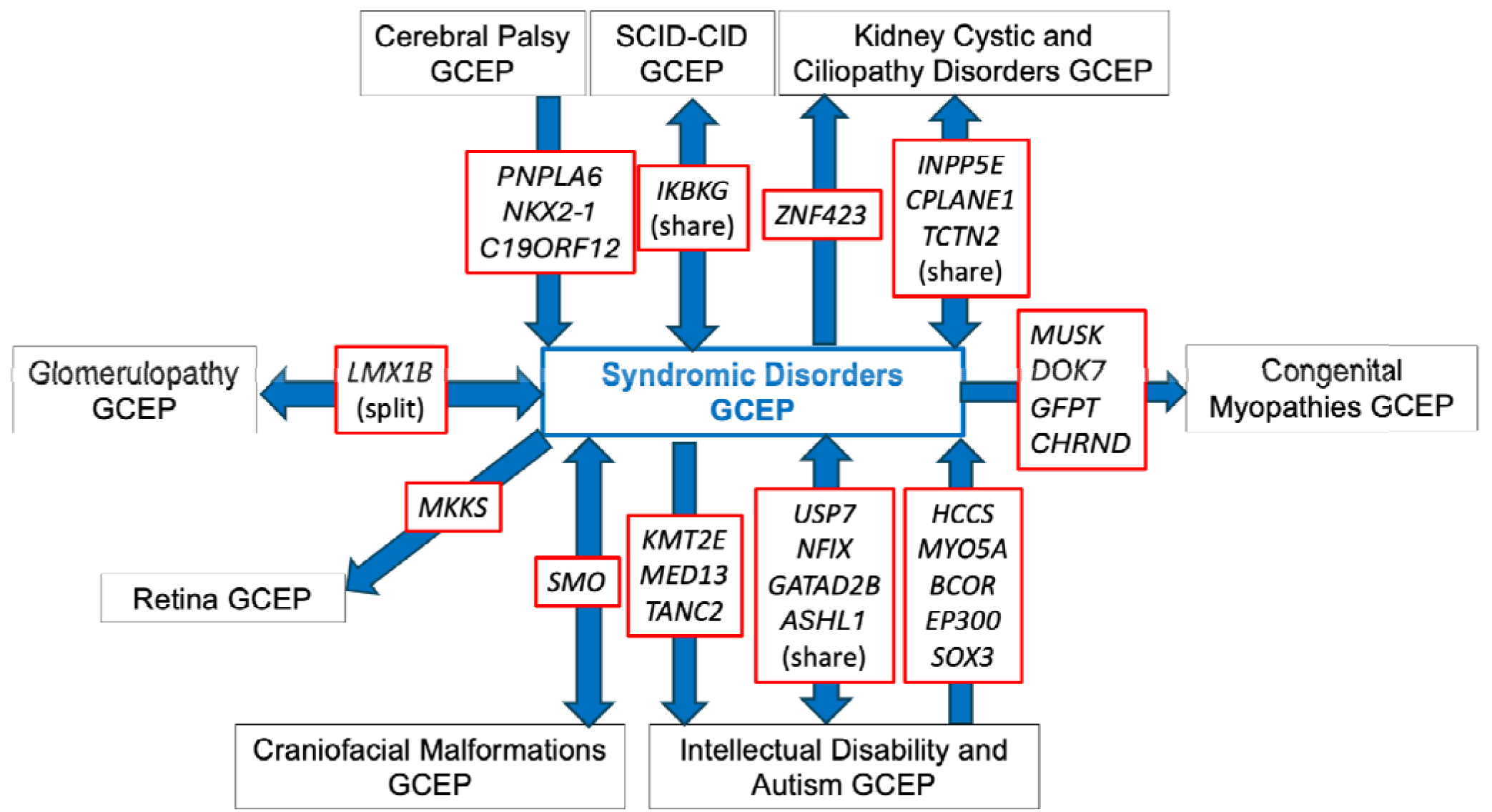
Exchange and collaboration on GDR curations between SD-GCEP and other GCEPs to date.

## Discussion

The SD-GCEP was created to address a gap in evaluating the clinical validity of GDRs involving multiple body systems and not under the purview of existing GCEPs. Additionally, the work of the SD-GCEP highlights the strength of the ClinGen framework. Not every expert has expertise in every syndrome, but the efforts of these “expert generalists” combined with the ClinGen Framework led to consistent gene-disease classifications across a wide range of conditions. Accurate GDR classifications are necessary for reporting, since ACMG recommends that diagnostic gene panels include all GDRs that meet criteria for Definitive, Strong, or Moderate evidence as defined by ClinGen.^6^

The SD-GCEP also highlights the importance of collaborating with other GCEPs. These collaborations are necessary for the best-informed gene-disease classifications. For example, the Craniofacial Malformations GCEP performed a precuration on the gene *SMO* and recommended keeping the disease assertions for Curry-Jones syndrome and Pallister-Hall-like syndrome as split. Curry-Jones syndrome is a syndromic craniosynostosis, so the Craniofacial Malformations GCEP curated that assertion, but referred the Pallister-Hall-like syndrome assertion to be curated under the expertise of the SD-GCEP since it involves multiple body systems. Similarly, the SD-GCEP forwarded the *MKKS* gene to be curated under the expertise of the Retina GCEP due to the retinal phenotypes present. The SD-GCEP has also collaborated with the Primary Immune Regulatory Disorders (PIRD) GCEP, where the PIRD GCEP performed a secondary curation of *RAB27A* and Griscelli syndrome type 2, a rare autosomal recessive disease characterized by cutaneous hypopigmentation, immunodeficiency, and hemophagocytic lymphohistiocytosis^31^, adding additional phenotypic information on immunodeficiency to the evidence summary. Effective communication and collaboration between GCEPs is optimal for accurate gene-disease classifications.

Lumping multiple disease assertions after a precuration adds to the challenge of naming the curated disease entity. Rare diseases, including many syndromic disorders, are commonly named after clinicians and patients. This approach does not provide information about the expected clinical features as a result of variation within the gene of interest. The SD-GCEP follows dyadic naming convention as defined by the *ClinGen’s guidance and recommendations for monogenic disease nomenclature* for GDRs that reach a classification of Moderate, Strong, or Definitive.^16^ The new label for the entity contains the HGNC gene symbol with the related phenotype. Out of the 14 lumped curations, seven have required the creation of a new name and identifier for the disease entity. These discussions are held in collaboration with relevant ClinGen expert panels, and OMIM and Mondo nomenclature expert members of the SD-GCEP. For example, there were six disease assertions associated with the gene *PNPLA6*. During the precuration, the GCEP decided to lump the assertions cerebellar ataxia and spastic paraplegia into the term “*PNPLA6*-related spastic paraplegia with or without ataxia” and lump the terms Boucher-Neuhauser syndrome, Gordon Holmes syndrome, Laurence-Moon syndrome, and Oliver-McFarlane syndrome into the term “retinal dystrophy-ataxia-pituitary hormone abnormality-hypogonadism syndrome.” The GCEP worked closely with Mondo to have these new terms created (MONDO:0100149, MONDO:0100155).

Conversely, splitting multiple disease assertions during the precuration phase poses a challenge at the variant curation level. Gene curation and variant curation are closely connected because the final clinical validity classification can impact the expected variant classification.

One way that the GDR classification is impacted is through splitting disease assertions into separate entities based on the present data. For the 38 genes associated with multiple conditions precurated by the SD-GCEP, 66% (25/38) have been curated as split entities with two or more separate entities. For example, the *ENPP1* gene has three gene-disease assertions for arterial calcification generalized of infancy 1; hypophosphatemic rickets autosomal recessive 2; and hypopigmentation-punctate palmoplantar keratoderma syndrome or Cole disease. A precuration identified that biallelic loss-of-function is the underlying mechanism of pathogenicity for all entities. However, based on the differences in inheritance pattern and clinical phenotype found in hypopigmentation-punctate palmoplantar keratoderma syndrome, the SD-GCEP decided to split this entity and lump arterial calcification generalized of infancy 1, and hypophosphatemic rickets autosomal recessive 2 as one disease entity. The final clinical validity classification for the lumped entity was Definitive while the split entity only reached Limited classification. Although splitting is justified utilizing a framework that examines the molecular mechanism, phenotypic spectrum, and inheritance patterns of the asserted disease entities, it may result in a reduction in the final evidence scores because the case reports and functional evidence are restricted to a specific condition rather than a broader “lumped” entity.^15^ If the final classification is Limited, No evidence, Disputed, or Refuted, it can reflect a lack of sufficient evidence supporting the syndrome and therefore potentially reducing the diagnosis rate.^6^ The SD-GCEP recognizes this limitation and plans to reconcile the problem by reevaluating these GDRs as defined by the ClinGen Standard Gene-Disease Relationship Recuration Procedure.^32^ Reevaluation of these curations every 2-3 years allows the consideration of new data and the possibility of upgrading the classification.

Moving forward, the ClinGen SD-GCEP will continue to evaluate the clinical validity of GDRs, prioritizing genes listed as Strong or Definitive for a syndrome in GenCC and those recommended by our expert panel members. The GCEP will begin re-curating genes previously classified as Strong, Moderate, or Limited whilst continuing to collaborate with other relevant GCEPs. By defining the clinical validity of GDRs involved in syndromic disorders, the SD-GCEP enables the incorporation of more clinically relevant genes in genetic testing panels and provides a critical resource to the community to improve diagnostic rates and patient outcomes.

## Data Availability

The data used in this publication are available in the Supplemental File and the online version of this article. The Clinical Genome Resource Syndromic Disorders Gene Curation Expert Panel makes all curations publicly available on the Clinical Genome Resource website (https://search.clinicalgenome.org/kb/gene-validity/).

## Author Contributions

Conceptualization: M.T.D., A.O.D.L., A.J.C. Formal analysis, Visualization: E.C.B., V.N.G., A.J.C. Funding acquisition: J.S.B., M.T.D., A.O.L., A.J.C. Writing-original draft: E.C.B., V.N.G., A.B.B., A.O.D.L., A.J.C. Data curation, Writing-review & editing: E.C.B., V.N.G., A.B.B., P.A., M.B.B., J.S.B., K.B., B.M.B., M.P.B., A.B., B.T.B., N.J.B., An.C., Ad.C., J.X.C., M.C., A.R.C., M.T.D., S.D., M.A.E., A.N.G., H.G., K.L.G., T.H., A.H., J.M.H., M.Y.H., S.S.J., S.K., A.K., AL.K., R.N.T.L., S.E.L., G.L., JY.L., A.M., H.R.M., B.M., S.M., S.A.M., E.H.O., E.E.P. B.C.P., M.J.P., R.R., J.C.R., F.L.R., B.D.R.R.A., S.A.Sa., Z.Sc., S.A.Sc., Z.St., S.P.S., J.P.T., C.T., D.L.T., S.T., K.T., N.A.V., B.W., R.F.W., A.O.D.L., A.J.C.

## Acknowledgements

Thank you to Jennifer Goldstein for her very valuable contribution and input into the early stages of establishing the SD-GCEP.

