## Supplemental Tables for "The ClinGen Syndromic Disorders Gene Curation Expert Panel: Assessing the Clinical Validity of 111 Gene-Disease Relationships"

| **Gene** | **Disease Assertions** | **Approach** | **Lumping and Splitting Outcome** | **Approval Date** |
| --- | --- | --- | --- | --- |
| *CACNA1G* | Spinocerebellar ataxia 42, and Spinocerebellar ataxia 42, early-onset, severe, with neurodevelopmental deficits | 2 | Lump | June 19^th^, 2020 |
| *BCOR* | Microphthalmia, syndromic 2 (OMIM:300166), Oculofaciocardiodental (OFCD) | 2 | Lump | August 5^th^, 2020 |
| *ESCO2* | Roberts-SC phocomelia syndrome (MONDO:0100253) | 1 | Lump | January 15^th^, 2021 |
| *LMX1B* | Focal segmental glomerulosclerosis 10 (OMIM:256020) and Nail-patella syndrome (OMIM:161200) | 1 | Split | February 3^rd^, 2021 |
| *WASHC5* | Ritscher-Schinzel syndrome 1 (OMIM: 220210) and hereditary spastic paraplegia 8 (OMIM: 603563) | 1 | Split | March 19^th^, 2021 |
| *WDR35* | Cranioectodermal dysplasia 2 (OMIM: 613610), and Short-rib thoracic dysplasia 7 with or without polydactyly (OMIM:614091) | 1 | Split | April 7^th^, 2021 |
| *ZMPSTE24* | mandibuloacral dysplasia with type B lipodystrophy (OMIM:608612), autosomal recessive lethal restrictive dermopathy (OMIM:275210), and Hutchinson-Gilford progeria syndrome | 1 | Split | April 7^th^, 2021 |
| *EP300* | Rubinstein-Taybi syndrome 2 (OMIM:613684), and Menke-Hennekam syndrome 2 (OMIM:618333) | 2 | Split | April 16^th^, 2021 |
| *ATP1A3* | Dystonia 12 (OMIM: 128235), Alternating hemiplegia of childhood 2 (OMIM:614820), developmental and epileptic encephalopathy 99 (OMIM:619606), and Cerebellar Ataxia, Areflexia, Pes Cavus, Optic Atrophy, And Sensorineural Hearing Loss (CAPOS) (OMIM:601338) | 2 | Lump | April 27^th^, 2021 |
| *UNC13A* | congenital nervous system disorder | 2 | Lump | June 18^th^, 2021 |
| *WNT10A* | Schopf-Schulz-Passarge syndrome (OMIM:224750), Tooth agenesis, selective, 4 (OMIM:150400), and Odontoonychodermal dysplasia (OMIM:257980) | 1 | Lump | July 16^th^, 2021 |
| *CENPJ* | microcephaly 6, primary, autosomal recessive (OMIM: 608393) and Seckel syndrome 4 (OMIM: 613676) | 1 | Lump | October 15^th^, 2021 |
| *CPLANE1* | Joubert syndrome 17(OMIM: 614615), and Orofaciodigital syndrome VI (OMIM: 277170) | 2 | Lump | October 26^th^, 2021 |
| *HSPG2* | Dyssegmental dysplasia, Silverman-Handmaker type(OMIM: 224410), and Schwartz-Jampel syndrome, type 1 (OMIM:255800) | 1 | Split | October 26^th^, 2021 |
| *SMCHD1* | Bosma-arhinia-microphthalmia-syndrome or BAMS (OMIM: 603457), and facioscapulohumeral muscular dystrophy 2 or FSHD2 (OMIM: 158901) | 2 | Split | November 3^rd^, 2021 |
| *INPP5E* | Impaired intellectual development, truncal obesity, retinal dystrophy, and micropenis syndrome (OMIM:610156), and Joubert syndrome 1 (OMIM:213300) | 4 | Split | December 17^th^, 2021 |
| *NFIX* | Malan syndrome (OMIM:614753), and Marshall-Smith syndrome (OMIM:602535) | 4 | Split | January 21^st^ , 2022 |
| *IKBKG* | Incontinentia pigmenti(OMIM:308300), Autoinflammatory disease, systemic, X-linked (OMIM:301081), Immunodeficiency 33 (OMIM:300636), and Ectodermal dysplasia and immunodeficiency 1 (OMIM:300291) | 4 | Split | March 2^nd^, 2022 |
| *ENPP1* | Cole disease(OMIM:615522), Hypophosphatemic rickets, autosomal recessive, 2(OMIM:613312), Arterial calcification, generalized, of infancy, 1 (OMIM:208000), {Diabetes mellitus, non-insulin-dependent, susceptibility to} (OMIM:125853), and {Obesity, susceptibility to} (OMIM:601665) | 3 | Split | March 18^th^, 2022 |
| *NKX2-1* | {Thyroid cancer, nonmedullary, 1} (OMIM:188550), Chorea, hereditary benign (OMIM:118700), and Choreoathetosis, hypothyroidism, and neonatal respiratory distress (OMIM: 610978) | 4 | Lump | April 6^th^, 2022 |
| *TREX1* | {Systemic lupus erythematosus, susceptibility to} (OMIM:152700), Aicardi-Goutieres syndrome 1, dominant and recessive (OMIM:225750), Chilblain lupus (OMIM: 610448), and Vasculopathy, retinal, with cerebral leukoencephalopathy and systemic manifestations (OMIM:192315) | 4 | Split | April 6^th^, 2022 |
| *NAA10* | Microphthalmia, syndromic 1(OMIM:309800),and Ogden syndrome (OMIM:300855) | 1 | Split | May 20^th^, 2022 |
| *TCTN2* | Joubert syndrome 24 (OMIM:616654), and ?Meckel syndrome 8 (OMIM:613885) | 1 | Lump | June 1^st^, 2022 |
| *DDR2* | Spondylometaepiphyseal dysplasia, short limb-hand type (OMIM:271665), and Warburg-Cinotti syndrome (OMIM:618175) | 2 | Split | July 6^th^, 2022 |
| *ZNF423* | Joubert syndrome 19(OMIM:614844), and Nephronophthisis 14 (OMIM:614844) | 4 | Lump; transferred to KCCD GCEP | July 6^th^, 2022 |
| *ERCC6* | Cockayne spectrum with or without cerebrooculofacioskeletal syndrome (OMIM: 133540, 214150, 278800), premature ovarian failure 11 (OMIM: 616946), UV-sensitive syndrome 1 (OMIM: 600630), and susceptibilities to [lung] cancer (OMIM: 211980) and age-related macular degeneration (OMIM: 613761) | 3 | Split | July 15^th,^ 2022 |
| *ERCC8* | Cockayne syndrome type 1 (OMIM: 216400) and UV-sensitive syndrome 2 (OMIM: 614621) | 3 | Split | July 15^th^, 2022 |
| *PNPLA6* | Boucher-Neuhauser syndrome (OMIM:215470), Oliver-McFarlane syndrome (OMIM:275400), Laurence-Moon syndrome (OMIM:245800), Gordon-Holmes syndrome (Orphanet lists this as cerebellar ataxia hypogonadism syndrome (ORPHA:1173), and spastic paraplegia 39, autosomal recessive (OMIM:612020) | 1 | Split | September 16^th^, 2022 |
| *C19ORF12* | Neurodegeneration with brain iron accumulation 4 (OMIM:614298), ?Spastic paraplegia 43, autosomal recessive (OMIM:615043) | 2 | Split | November 22^nd^, 2022 |
| *LRP4* | ?Myasthenic syndrome, congenital, 17 (OMIM:616304), Cenani-Lenz syndactyly syndrome (OMIM:212780), and Sclerosteosis 2 (OMIM:614305) | 1 | Split | March 17^th^, 2023 |
| *RARB* | Microphthalmia, syndromic 12 (OMIM:615524) | 1 | Lump | March 17^th^, 2023 |
| *PLCB4* | Auriculocondylar syndrome 2A (OMIM:614669) and Auriculocondylar syndrome 2B (OMIM:620458) | 2 | Split | Wednesday, April 5^th^, 2023 |
| *NOG* | Brachydactyly, type B2 (OMIM:611377), Multiple synostoses syndrome 1 (OMIM:186500), Stapes ankylosis with broad thumbs and toes (OMIM:184460), Symphalangism, proximal, 1A (OMIM:185800), and Tarsal-carpal coalition syndrome (OMIM:186570) | 1 | Lump | May 19^th^, 2023 |
| *SOX3* | panhypopituitarism, X-linked (OMIM: 312000) and intellectual developmental disorder, X-linked, with isolated growth hormone deficiency (OMIM: 300123), 46,XX sex reversal 3 (MONDO:0010442), X-linked congenital generalized hypertrichosis (ORPHA:79495) and Septo-optic dysplasia spectrum (ORPHA:3157) | 4 | Split | June 16^th^, 2023 |
| *MAB21L2* | Microphthalmia/coloboma and skeletal dysplasia syndrome (OMIM:615877) | 1 | Split | July 25^th^, 2023 |
| *TET3* | Beck-Fahrner syndrome (OMIM:618798) | 2 | Split | November 1^st^, 2023 |
| *NARS1* | autosomal dominant (AD) neurodevelopmental disorder with microcephaly, impaired language, epilepsy, and gait abnormalities (NEDMILEG) (OMIM:619092) and autosomal recessive (AR) neurodevelopmental disorder with microcephaly, impaired language, and gait abnormalities (OMIM:619091) | 3 | Split | February 7t^h^, 2024 |
| *MKKS* | Bardet-Biedl syndrome 6 (OMIM:605231), and McKusick-Kaufman syndrome (OMIM:236700) | 4 | Lump; transferred to Retina GCEP | February 16^th^, 2024 |

**Supplementary Table 1:** **Precurations outcomes performed by the SD-GCEP.** Approach 1: GDRs most frequently tested in clinical laboratories, Approach 2: GDRs through clinical genome or exome sequencing performed by diagnostic laboratories within the membership of the SD-GCEP, Approach 3: new GDRs from research consortia including NHGRI’s Centers of Mendelian Genomics and GREGoR consortium, Approach 4: GDRs requested by other GCEPs for phenotypes requiring the broad expertise of the SD-GCEP, Approach 5: syndromic GDRs in GenCC with Strong or Definitive classifications not curated by other GCEPs.

| **Gene** | **Disease Assertion** | **MOI** | **Approach** | **Classification** | **Approval Date** |
| --- | --- | --- | --- | --- | --- |
| *ARL13B* | Joubert syndrome (MONDO:0018772) | AR | 1 | Definitive | June 3^rd^, 2020 |
| *TSHZ1* | Aural atresia, congenital. Autosomal dominant inheritance (MONDO:0011921) | AD | 2 | Limited | June 19^th^, 2020 |
| *ZNF462* | Weiss-Kruszka syndrome (MONDO:0032836) | AD | 2 | Definitive | July 17^th^, 2020 |
| *ARSL* | X-linked chondrodysplasia punctata 1 (MONDO:0010555) | XLR | 1 | Definitive | July 28^th^, 2020 |
| *TCF20* | developmental delay with variable intellectual impairment and behavioral abnormalities (MONDO:0032745) | AD | 2 | Definitive | July 28^th^, 2020 |
| *POLR2A* | neurodevelopmental disorder with hypotonia and variable intellectual and behavioral abnormalities, autosomal dominant (MONDO:0032829) | AD | 3 | Moderate | August 5^th^, 2020 |
| *NOD2* | Blau syndrome (MONDO:0008523) | AD | 2 | Definitive | August 21^st^, 2020 |
| *TMEM237* | Joubert syndrome 14 (MONDO:0013745) | AR | 1 | Definitive | August 21^st^, 2020 |
| *LIFR* | Stüve-Wiedemann syndrome (MONDO:0011108) | AR | 1 | Definitive | September 2^nd^, 2020 |
| *BCOR* | microphthalmia, syndromic 2 (MONDO:0010261) | XLD | 2 | Definitive | September 18^th^, 2020 |
| *PAX7* | congenital myopathy with myasthenic-like onset (MONDO:0018528) | AR | 2 | Moderate | September 18^th^, 2020 |
| *PPP1R12A* | genitourinary and/or brain malformation syndrome (MONDO:0032934) | AD | 2 | Strong | October 16^th^, 2020 |
| *RAB27A* | Griscelli syndrome type 2 (MONDO:0011872) | AR | 1 | Definitive | October 27^th^, 2020 |
| *WNT5A* | Robinow syndrome, autosomal dominant (MONDO:0008389) | AD | 2 | Moderate | October 27^th^, 2020 |
| *UNC80* | hypotonia-speech impairment-severe cognitive delay syndrome (MONDO:0018297) | AR | 2 | Definitive | November 4^th^, 2020 |
| *USP7* | Hao-Fountain syndrome (MONDO:0014805) | AD | 2 | Definitive | December 2^nd^, 2020 |
| *DCAF17* | Woodhouse-Sakati syndrome (MONDO:0009419) | AR | 1 | Definitive | December 18^th^, 2020 |
| *CAMK2G* | intellectual developmental disorder 59 (MONDO:0032795) | AD | 2 | Limited | January 6^th^, 2021 |
| *UBE3B* | oculocerebrofacial syndrome, Kaufman type (MONDO:0009485) | AR | 2 | Definitive | January 6^th^, 2021 |
| *MYO5A* | Griscelli syndrome type 1 (MONDO:0008962) | AR | 2 | Definitive | January 26^th^, 2021 |
| *TBX5* | Holt-Oram syndrome (MONDO:0007732) | AD | 1 | Definitive | January 26^th^, 2021 |
| *OGT* | intellectual disability, X-linked 106 (MONDO:0030907) | XLR | 3 | Moderate | February 3^rd^, 2021 |
| *LMX1B* | nail-patella syndrome (MONDO:0008061) | AD | 1 | Definitive | February 3^rd^, 2021 |
| *SMARCAL1* | Schimke immuno-osseous dysplasia (MONDO:0009458) | AR | 1 | Definitive | February 19^th^, 2021 |
| *ESCO2* | Roberts-SC phocomelia syndrome (MONDO:0100253) | AR | 1 | Definitive | March 19^th^, 2021 |
| *PRG4* | camptodactyly-arthropathy-coxa vara-pericarditis syndrome (MONDO:0008828) | AR | 2 | Definitive | April 27^th^, 2021 |
| *ZMPSTE24* | Mandibuloacral dysplasia with type B lipodystrophy (MONDO:0012074) | AR | 1 | Definitive | June 2^nd^, 2021 |
| *ZMPSTE24* | Restrictive dermopathy (MONDO:0010143) | AR | 1 | Definitive | June 2^nd^, 2021 |
| *IFT122* | cranioectodermal dysplasia 1 (MONDO:0021093) | AR | 1 | Definitive | June 18^th^, 2021 |
| *ATP1A3* | ATP1A3-related neurological disorders (MONDO:0700002) | AD | 2 | Definitive | July 16^th^, 2021 |
| *WDR35* | cranioectodermal dysplasia (MONDO:0013323) | AR | 1 | Definitive | August 4^th^, 2021 |
| *WDR35* | short-rib thoracic dysplasia 7 with or without polydactyly (MONDO:0013569) | AR | 1 | Definitive | August 4^th^, 2021 |
| *PCGF2* | Turnpenny-Fry syndrome (MONDO:0032707) | AD | 3 | Strong | August 20^th^, 2021 |
| *WNT10A* | ectodermal dysplasia WNT10A related (MONDO:0100358) | AR | 1 | Definitive | August 20^th^, 2021 |
| *EP300* | Rubinstein-Taybi syndrome due to EP300 haploinsufficiency (MONDO:0013364) | AD | 2 | Definitive | September 1^st^, 2021 |
| *WASHC5* | Ritscher-Schinzel syndrome 1 (MONDO:0009073) | AR | 1 | Limited | September 17^th^, 2021 |
| *WASHC5* | Spastic paraplegia 8 (MONDO:0011339) | AD | 1 | Moderate | September 17^th^, 2021 |
| *UNC13A* | congenital nervous system disorder (MONDO:0002320) | U | 2 | Limited  Provisional status: Awaiting additional genetic evidence. | October 15^th^, 2021 |
| *MAPK8IP3* | neurodevelopmental disorder with or without variable brain abnormalities; NEDBA (MONDO:0032755) | AD | 2 | Strong | October 26^th^, 2021 |
| *KMT5B* | complex neurodevelopmental disorder (MONDO:0011518) | AD | 2 | Definitive | November 3^rd^, 2021 |
| *SMCHD1* | arhinia, choanal atresia, and microphthalmia (MONDO:0011323) | DD | 2 | Definitive | November 3^rd^, 2021 |
| *SPTBN4* | neurodevelopmental disorder with hypotonia, neuropathy, and deafness (MONDO:0060496) | AR | 2 | Definitive | December 1^st^, 2021 |
| *CPLANE1* | Joubert syndrome 17 (MONDO:0013824) | AR | 2 | Definitive | December 17^th^, 2021 |
| *CENPJ* | autosomal recessive microcephaly 6 with or without short stature (MONDO:0700054) | AR | 1 | Definitive | January 5^th^, 2022 |
| *CSNK2B* | Poirier-Bienvenu neurodevelopmental syndrome (MONDO:0032889) | AD | 2 | Definitive | January 21^st^, 2022 |
| *KMT2A* | Wiedemann-Steiner syndrome (MONDO:0011518) | AD | 2 | Definitive | February 23^rd^, 2022 |
| *MYMK* | Carey-Fineman-Ziter syndrome (MONDO:0009700) | AR | 2 | Definitive | March 2^nd^, 2022 |
| *GATAD2B* | severe intellectual disability-poor language-strabismus-grimacing face-long fingers syndrome (MONDO:0014034) | AD | 2 | Definitive | March 18^th^, 2022 |
| *HSPG2* | Silverman-Handmaker type dyssegmenta dysplasia (MONDO:0009140) | AR | 1 | Definitive | April 26^th^, 2022 |
| *HSPG2* | Schwartz-Jampel syndrome type 1 (MONDO:0100435) | AR | 1 | Definitive | April 26^th^, 2022 |
| *PIGU* | Glycosylphosphatidylinositol biosynthesis defect 21 (MONDO:0032824) | AR | 3 | Limited | May 20^th^, 2022 |
| *IKBKG* | Incontinentia pigmenti (MONDO:0010631) | XLD | 4 | Definitive | June 1^st^, 2022 |
| *ENPP1* | Generalized arterial calcification of infancy (MONDO:0008817) | AR | 3 | Definitive | June 17^th^, 2022 |
| *ENPP1* | Hypopigmentation-punctate palmoplantar keratoderma syndrome (MONDO:0014227) | AD | 3 | Limited | June 17^th^, 2022 |
| *INPP5E* | MORM syndrome (MONDO:0012423) | AR | 4 | Moderate | July 26^th^, 2022 |
| *NKX2-1* | NKX2-1 related choreoathetosis and congenital hypothyroidism with or without pulmonary dysfunction (MONDO:0100520) | AD | 4 | Definitive | July 26^th^, 2022 |
| *NFIX* | Malan overgrowth syndrome (MONDO:0013885) | AD | 4 | Definitive | August 3^rd^, 2022 |
| *NFIX* | Marshall-Smith syndrome (MONDO:0011244) | AD | 4 | Definitive | August 3^rd^, 2022 |
| *ERCC6* | Cockayne spectrum with or without cerebrooculofacioskeletal syndrome (MONDO:0100506) | AR | 3 | Definitive | August 19^th^, 2022 |
| *ERCC8* | Cockayne syndrome type 1 (MONDO:0019569) | AR | 3 | Definitive | August 19^th^, 2022 |
| *CDC45* | Meier-Gorlin syndrome 7 (MONDO:0014894) | AR | 1 | Definitive | October 5^th^, 2022 |
| *DDR2* | Warburg-Cinotti syndrome (MONDO:0032579) | AD | 2 | Moderate | October 21^st^, 2022 |
| *DDR2* | Spondyloepimetaphyseal dysplasia-short limb-abnormal calcification syndrome (MONDO:0010077) | AR | 2 | Definitive | October 21^st^, 2022 |
| *PNPLA6* | PNPLA6-related spastic paraplegia with or without ataxia (MONDO:0100149) | AR | 1 | Definitive | November 2^nd^, 2022 |
| *PNPLA6* | Retinal dystrophy-ataxia-pituitary hormone abnormality-hypogonadism syndrome (MONDO:0100155) | AR | 1 | Definitive | November 2^nd^, 2022 |
| *GRIA2* | Neurodevelopmental disorder with language impairment and behavioral abnormalities (MONDO:0030060) | AD | 2 | Definitive | November 18^th^, 2022 |
| *PBX1* | Congenital anomalies of kidney and urinary tract syndrome with or without hearing loss, abnormal ears, or developmental delay (MONDO:0060549) | AD | 2 | Definitive | November 18^th^, 2022 |
| *ANTXR2* | Hyaline fibromatosis syndrome (MONDO:0009229) | AR | 1 | Definitive | November 22^nd^, 2022 |
| *CDT1* | Meier-Gorlin syndrome 4 (MONDO:0013431) | AR | 1 | Definitive | December 7^th^, 2022 |
| *GPAA1* | Glycosylphosphatidylinositol biosynthesis defect 15 (MONDO:0060627) | AR | 1 | Strong | December 7^th^, 2022 |
| *TCTN2* | Joubert syndrome 24 (MONDO:0014724) | AR | 1 | Definitive | February 1^st^, 2023 |
| *SH3PXD2B* | Frank-Ter Haar syndrome (MONDO:0009579) | AR | 1 | Definitive | February 1^st^, 2023 |
| *ORC4* | Meier-Gorlin syndrome 2 (MONDO:0013428) | AR | 1 | Moderate | February 17^th^, 2023 |
| *C19ORF12* | neurodegeneration with brain iron accumulation 4 (MONDO:0013674) | AR | 2 | Definitive | February 28^th^, 2022 |
| *C19ORF12* | neurodegeneration with brain iron accumulation 4 (MONDO:0013674) | AD | 2 | Moderate | February 28^th^, 2022 |
| *RPL5* | Diamond-Blackfan anemia 6 (MONDO:0012937) | AD | 1 | Definitive | March 1^st^, 2023 |
| *RPS19* | Diamond-Blackfan anemia (MONDO:0015253) | AD | 1 | Definitive | March 1^st^, 2023 |
| *NCAPG2* | Khan-Khan-Katsanis syndrome (MONDO:0032764) | AR | 2 | Limited | April 5^th^, 2023 |
| *LRP4* | congenital myasthenic syndrome 17 (MONDO:0014578) | AR | 1 | Limited | April 21^st^, 2023 |
| *LRP4* | Cenani-Lenz syndactyly syndrome (MONDO:0008931) | AR | 1 | Definitive | April 21^st^, 2023 |
| *RARB* | Microphthalmia, syndromic 12 (MONDO:0014229) | AD | 1 | Definitive | April 25^st^, 2023 |
| *PLCB4* | Auriculocondylar syndrome 2 (MONDO:0013845) | AD | 2 | Definitive | June 7^th^, 2023 |
| *PLCB4* | Auriculocondylar syndrome 2 (MONDO:0013845) | AR | 2 | Moderate | June 7^th^, 2023 |
| *NOG* | NOG-related symphalangism spectrum disorder (MONDO:0100521) | AD | 1 | Definitive | July 5^th^, 2023 |
| *ACTL6A* | ACTL6A-related BAFopathy (MONDO:0700121) | AD | 2 | Moderate | July 21^st^, 2023 |
| *ORC6* | Meier-Gorlin syndrome 3 (MONDO:0013430) | AR | 1 | Definitive | July 21^st^, 2023 |
| *SOX3* | X-linked intellectual disability with hypopituitarism (MONDO:0100195) | XL | 4 | Moderate | August 2^nd^, 2023 |
| *MAB21L2* | colobomatous microphthalmia-rhizomelic dysplasia syndrome (MONDO:0014380) | AD | 1 | Definitive | September 6^th^, 2023 |
| *GEMIN5* | Neurodevelopmental disorder with cerebellar atrophy and motor dysfunction (MONDO:0859152) | AR | 2 | Definitive | September 6^th^, 2023 |
| *NRROS* | Seizures, early-onset, with neurodegeneration and brain calcification (MONDO:0030033) | AR | 2 | Definitive | September 15^th^, 2023 |
| *AGO2* | Lessel-Kreienkamp syndrome (MONDO:0030897) | AD | 2 | Definitive | September 15^th^, 2023 |
| *NEMF* | intellectual developmental disorder with speech delay and axonal peripheral neuropathy (MONDO:0030849) | AR | 3 | Strong | October 4^th^, 2023 |
| *IGF2* | Silver-Russell syndrome 3 (MONDO:0014663) | AD | 2 | Definitive | October 4^th^, 2023 |
| *SMG8* | Alzahrani-Kuwahara syndrome (MONDO:0859136) | AR | 2 | Definitive | October 20^th^, 2023 |
| *SMO* | Pallister-Hall-like syndrome (MONDO:0009436) | AR | 4 | Moderate | October 20^th^, 2023 |
| *PPP1R21* | neurodevelopmental disorder with hypotonia, facial dysmorphism, and brain abnormalities (MONDO:0859165) | AR | 2 | Strong | October 24^th^, 2023 |
| *SPEN* | Radio-Tartaglia syndrome (MONDO:0859143) | AD | 3 | Definitive | November 1^st^, 2023 |
| *TET3* | Beck-Fahrner syndrome (MONDO:0032922) | AR | 2 | Limited | November 17^th^, 2023 |
| *TET3* | Beck-Fahrner syndrome (MONDO:0032922) | AD | 2 | Definitive | November 17^th^, 2023 |
| *TRAPPC4* | neurodevelopmental disorder with epilepsy, spasticity, and brain atrophy (MONDO:0032894) | AR | 3 | Definitive | December 6^th^, 2023 |
| *VARS1* | neurodevelopmental disorder with microcephaly, seizures, and cortical atrophy (MONDO:0060621) | AR | 2 | Definitive | December 6^th^, 2023 |
| *MED27* | neurodevelopmental disorder with spasticity, cataracts, and cerebellar hypoplasia (MONDO:0859137) | AR | 3 | Strong | December 15^th^, 2023 |
| *SPTBN1* | developmental delay, impaired speech, and behavioral abnormalities (MONDO:0859178) | AD | 2 | Strong | December 15^th^, 2023 |
| *PRR12* | neuroocular syndrome (MONDO:0859193) | AD | 2 | Definitive | January 19^th^, 2024 |
| *TXNL4A* | choanal atresia-hearing loss-cardiac defects-craniofacial dysmorphism syndrome (MONDO:0012064) | AR | 5 | Definitive | February 16^th^, 2024 |
| *NARS1* | neurodevelopmental disorder with microcephaly, impaired language, and gait abnormalities (MONDO:0100348) | AR | 3 | Moderate | March 6^th^, 2024 |
| *NARS1* | neurodevelopmental disorder with microcephaly, impaired language, epilepsy, and gait abnormalities (MONDO:0030837) | AD | 3 | Moderate | March 6^th^, 2024 |
| *DPH1* | developmental delay with short stature, dysmorphic facial features, and sparse hair (MONDO:0031632) | AR | 5 | Definitive | March 15^th^, 2024 |
| *THOC6* | THOC6-related developmental delay-microcephaly-facial dysmorphism syndrome (MONDO:0013362) | AR | 5 | Definitive | March 15^th^, 2024 |
| *H4C5* | Tessadori-Van Haaften neurodevelopmental syndrome 3 (MONDO:0030993) | AD | 5 | Strong | March 19^th^, 2024 |
| *H3-3A* | Bryant-Li-Bhoj neurodevelopmental syndrome 1 (MONDO:0030606) | AD | 2 | Definitive | March 19^th^, 2024 |

**Supplemental Table 2: Curation outcomes performed by the SD-GCEP.** MOI: Mode of inheritance. Approach 1: GDRs most frequently tested in clinical laboratories, Approach 2: GDRs through clinical genome or exome sequencing performed by diagnostic laboratories within the membership of the SD-GCEP, Approach 3: new GDRs from research consortia including NHGRI’s Centers of Mendelian Genomics and GREGoR consortium, Approach 4: GDRs requested by other GCEPs for phenotypes requiring the broad expertise of the SD-GCEP, Approach 5: syndromic GDRs in GenCC with Strong or Definitive classifications not curated by other GCEPs.
